# Five-Year Survival Outcomes for Breast Cancer Patients Across Continental Africa: A Contemporary Review of Literature with Meta Analysis

**DOI:** 10.1101/2025.01.03.25319952

**Authors:** Augustina Badu-Peprah, Ernest Kissi Kontor, Adu-Gyamfi Benjamin, Jessica Kumah, Akosua Aya Essuman, Bossoh Selorm, Issahak Nurudeen, Bismark Osei Owusu, Nityanand Jain

**Author notes:** Co-first authors (contributed equally) (Augustina Badu-Peprah, Ernest Kissi Kontor).

## Abstract

**Background:** Breast cancer associated mortality in Africa remains high due to poor survival rates, varying widely across countries. Despite medical advancements, barriers like limited access to early detection and treatment persist. This meta-analysis offers a crucial update on 5-year survival trends and influencing factors across continental Africa.

**Methods:** A systematic search of four biomedical databases and citation searching identified 79 articles from 22 African countries, analyzing 27,559 patients (97% female). A random-effects model was used to estimate the 5-year survival rate with subgroup analyses. Publication bias was assessed using Egger’s test and funnel plots.

**Results:** Pooled overall 5-year breast cancer survival in Africa averaged 48% (95% CI: 43-53%) with high statistical heterogeneity (I² = 98%). Survival was highest in Northern Africa (64%; 95% CI: 59-69%) and lowest in Western Africa (32%; 95% CI: 23-42%). Males (51%; 95% CI: 36-65%) had marginally higher survival than females (48%; 95% CI: 42-54%). Socioeconomic indices were positively associated with better outcomes. Publication bias, adjusted by the trim-and-fill method, raised survival to 62% (95% CI: 55-67%). A country-wise comparison with 2018 estimates suggests a declining survival tendency, with WHO AFRO countries reporting the poorest estimates among other WHO regions. Despite regional differences, survival trends seem to have plateaued near 48-49% level continent-wide since the early 2010s.

**Conclusions:** Our findings reveal marked regional disparities in survival rates across Africa, underscoring the urgent need for targeted healthcare interventions. Strengthening healthcare systems, ensuring universal access, and driving socioeconomic progress are vital to improving survival outcomes.

## 1. Introduction

Breast cancer remains a significant global health concern that affects both women and men, despite the considerable progress made in early diagnostics and treatment modalities [1]. It is the most prevalent malignant disease among women worldwide and is the second leading cause of cancer-related mortality [2,3]. The heterogeneity and complexity in managing breast cancer is underscored by its multiple subtypes, which exhibit varying biological behaviors and clinical responses to treatment [4,5]. Notwithstanding substantial advancements in early detection and therapeutic strategies, significant survival disparities persist between high-income and low-to middle-income countries [6,7].

In Africa, the burden of breast cancer is particularly pronounced, with incidence and mortality rates continuing to rise. The latest estimates indicate that breast cancer leads to an estimated 186,598 new cases and 85,787 deaths annually among Africans [8]. Notably, regional variations in disease burden have been observed within the continent, with the highest incidence of new cases occurring in Northern and Western Africa [9]. Contributing factors include delayed diagnosis, inadequate healthcare infrastructure and funding, insufficient public health education initiatives, mistrust in western medicine, and significant socioeconomic barriers.

The most recent 5-year survival estimates for African countries were reported by a meta-analysis conducted by Ssentongo et al. [10]. Their study, which was limited to a search strategy till October 2018, determined that the overall 5-year survival rate in Africa was 52.9%. Subsequently, no further estimates have been collated, potentially resulting in a discrepancy between the literature and the situation on the ground. Cochrane suggests that updates should occur biennially, while Campbell reviews recommend that updates occur within a five-year cycle to facilitate the accumulation of knowledge [11].

These updates are driven by the rapid evolution of medical research and treatment methodologies, which may not align with current practices, advancements, or emerging trends observed six years ago. Additionally, the socioeconomic and healthcare landscapes in Africa are subject to continual change influenced by several factors, including policies, investments, and regional partnerships. These dynamics can significantly impact breast cancer diagnosis, treatment, and survival outcomes, highlighting the necessity for an updated analysis.

Hence, we believe that the integration of more recent data allows for a detailed and updated analysis of survival outcomes in Africa. The findings from our study will provide insights that can be acted upon, offering a detailed understanding of the factors influencing breast cancer survival across diverse African contexts. Such information is crucial for the development of targeted interventions, the shaping of policy decisions, and ultimately the improvement of patient outcomes across the region.

## 2. Methods

The protocol for the present systematic review and meta-analysis was registered prospectively on PROSPERO (registration number – CRD42024595654). The Preferred Reporting Items for Systematic Reviews and Meta-Analyses (PRISMA) guidelines were followed throughout the search and study selection process to ensure transparency and completeness. Discrepancies between the registered protocol and research methodology are reported in Appendix 1.

### 2.1. Data Sources and Search Strategy

A literature search was undertaken across multiple biomedical databases, including PubMed, Medline, Web of Science, and Scopus, to identify studies reporting 5-year survival estimates for breast cancer across African countries. The search was restricted to articles published from database inception to October 2024 to ensure the inclusion of the most recent data. We used the SPIDER framework (Sample, Phenomenon of Interest, Design, Evaluation, Research Type) to structure our search strategy as follows:

***Sample (S)***: Patients diagnosed with breast cancer in Africa.

***Phenomenon of Interest (PI):*** 5-year survival estimates.

***Design (D):*** Observational studies, retrospective and prospective.

***Evaluation (E):*** 5-year survival rates and socio-demographic factors influencing survival.

***Research Type (R):*** Quantitative studies that report survival rates.

We used a combination of author keywords and Medical Subject Headings (MeSH) terms such as “breast cancer,” “five-year survival,” “Africa,” “Kaplan Meier,” and “survival.” To enhance the comprehensiveness of our search, we performed a citation search by examining the reference lists of all relevant studies and published systematic reviews and meta-analyses (**Appendix 2**). Given that literature from low-resource settings tends to be reported in non-indexed gray literature, we non-comprehensively and manually searched Google Scholar. The search comprised of the string – *(“survival”) AND (“breast cancer”) AND (“name of country”)* and restricted to availability of these terms in the title of the paper. This was done for all 54 countries in Africa to ensure that no data is missed from countries that were not included in previous meta-analyses. No language restrictions were applied, but studies had to include an English abstract.

### 2.2. Study Selection

Studies were imported into the Covidence software and duplicates were automatically removed (except Google Scholar searches). EKK, AAE, JK, and SB independently screened the titles and abstracts of all identified articles. Full-text articles were then retrieved for studies that appeared to meet the inclusion criteria based on their abstracts. Studies were included if they reported 5-year survival rates for breast cancer patients, were conducted within African countries, provided sufficient data to calculate proportions, and were published in any language. Studies that did not provide separate survival rates for breast cancer, or were editorials, commentaries, randomized control trials or reviews without original data, were excluded. Conference abstracts were included if no corresponding full-text paper was found. For full-text screening, PDFs of the non-English articles were uploaded and translated using PDFSimpli (https://pdfsimpli.com/pdf-editor/translate-pdf/; accessed 09th November 2024).

Each study was carefully evaluated against the inclusion and exclusion criteria. Any discrepancies between reviewers during the study selection process were resolved through discussion with ABP and BAG. If consensus could not be reached, a third reviewer, NJ was consulted to make the final decision. The study selection process was meticulously documented using a PRISMA flow diagram to illustrate the number of records identified, screened, and assessed for eligibility and inclusion. For Google Scholar searches, results were not considered for inclusion in PRISMA flow diagram, primarily due to the non-comprehensive nature of our search and unavailability of results for majority of countries. For countries with results, the full texts were directly screened by NJ and EKK against the inclusion criteria and the final list of included studies from other databases. No new studies were identified from Google Scholar.

### 2.3. Data Extraction

Data from eligible studies were independently extracted by two reviewers using a standardized data extraction Excel form. Extraction was done in pairs by EKK and SB, ABP and BAG, and AAE and JK. Extracted data included study characteristics such as authors, year of publication, country, gender, region, race, median age, sample size, overall survival (OS), and number of participants alive at the end of 5-year follow-up period (n). The country’s Human Development Index (HDI) values was classified according to its HDI ranking in 2022 [12]. Socio-demographic Index (SDI), a measure from the Global Burden of Disease collaboration, was categorized into quantiles to provide a comprehensive understanding of the demographic context [13]. We also extracted the survival rates using Webplotdigitizer v.4.5., following the method described by Tierney et al. [14].

To account for sex differences in our analysis, we categorized studies with mixed male and female participants based on a threshold criterion. Specifically, studies were classified under a specific sex group if one sex constituted at least 80% of the sample population. This cutoff was selected to ensure that the overall survival outcomes would be predominantly driven by the characteristics of the majority sex group, thereby reducing potential confounding effects from mixed-sex samples. This method, though implicitly employed by other studies [10,15], validates its use in our work. Furthermore, we believe that such a methodological approach allows for a more accurate representation of the primary variable of interest in the sex-specific survival analysis. When necessary, corresponding authors were contacted to obtain additional information or clarifications. Any discrepancies in data extraction were resolved through consultation with a third reviewer, NJ. The extracted data were then reviewed and validated by the research team to ensure accuracy and consistency.

### 2.4. Quality Assessment

The quality assessment of the included studies was adapted from the methodology outlined by Ssentongo et al. (**Appendix 3**) [10]. This method was selected for its specificity to breast cancer meta-analysis and its ability to provide numerical outcomes that can be used to assess study-level covariates. The approach evaluates studies based on three primary criteria – detailed demographic information, stratified reporting of survival, and sample size. Each criterion has a maximum score, with demographic information scoring up to 11 points, stratified reporting of survival up to 7 points, and sample size up to 6 points. The higher the number of points, the better the quality of the assessed study.

### 2.5. Data Analysis

The survival rates were pooled using the random effects inverse variance method, employing the “*metaprop*” function in the <*meta*> package of R v.4.4.0. Proportions were transformed using the logit transformation, and the 95% confidence interval (CI) was based on the Clopper-Pearson interval (exact binomial interval). To estimate heterogeneity variance, we used the Restricted Maximum Likelihood (REML) method, which is more suitable for sub-groups involving a small number of studies, as the DerSimonian method can be misleading [16,17]. Subgroup analyses were conducted based on geographic region (Northern, Eastern, Central, Western, Southern Africa), patient sex and ethnicity, Human Development Index (HDI), Social Development Index (SDI), individual countries, and year of publication.

Statistical heterogeneity was assessed using the I² statistic and Cochran’s Q test. Publication bias was evaluated with funnel plots and Egger’s test, with adjustments made using the fill and trim method. Additionally, a limit meta-analysis was conducted. Meta-regression analyses examined the influence of study-level covariates on survival outcomes. The <*metafor*> and <*orchard*> packages were used for the meta-regression. A cumulative meta-analysis was conducted to assess changes in survival rates in Africa over time. We compared our results to previously reported estimates for Africa and other World Health Organization (WHO) regions using one-sample Z test.

## 3. Results

A total of 3884 records were identified through a comprehensive search of four biomedical databases and citation searching. After a thorough screening of abstracts, titles, and full texts, 79 records were deemed eligible for inclusion (**Figure 1**) [18–96]. As some records presented data for multiple countries or demographics within a single record, we designated a reference as a “record”, while each corresponding data was termed as a “study”. Accordingly, we included 88 studies in the pooled meta-analysis. Similarly, 91 studies were analyzed for sex-specific survival outcomes. Data from 27,559 individuals was pooled in our study, with a female-to-male ratio of 26,709 to 850 (97% to 3%), respectively. The articles were sourced from 22 of the 54 countries in Africa. Study and sampling characteristics are summarized in **Appendix 4**.

**Figure 1.**
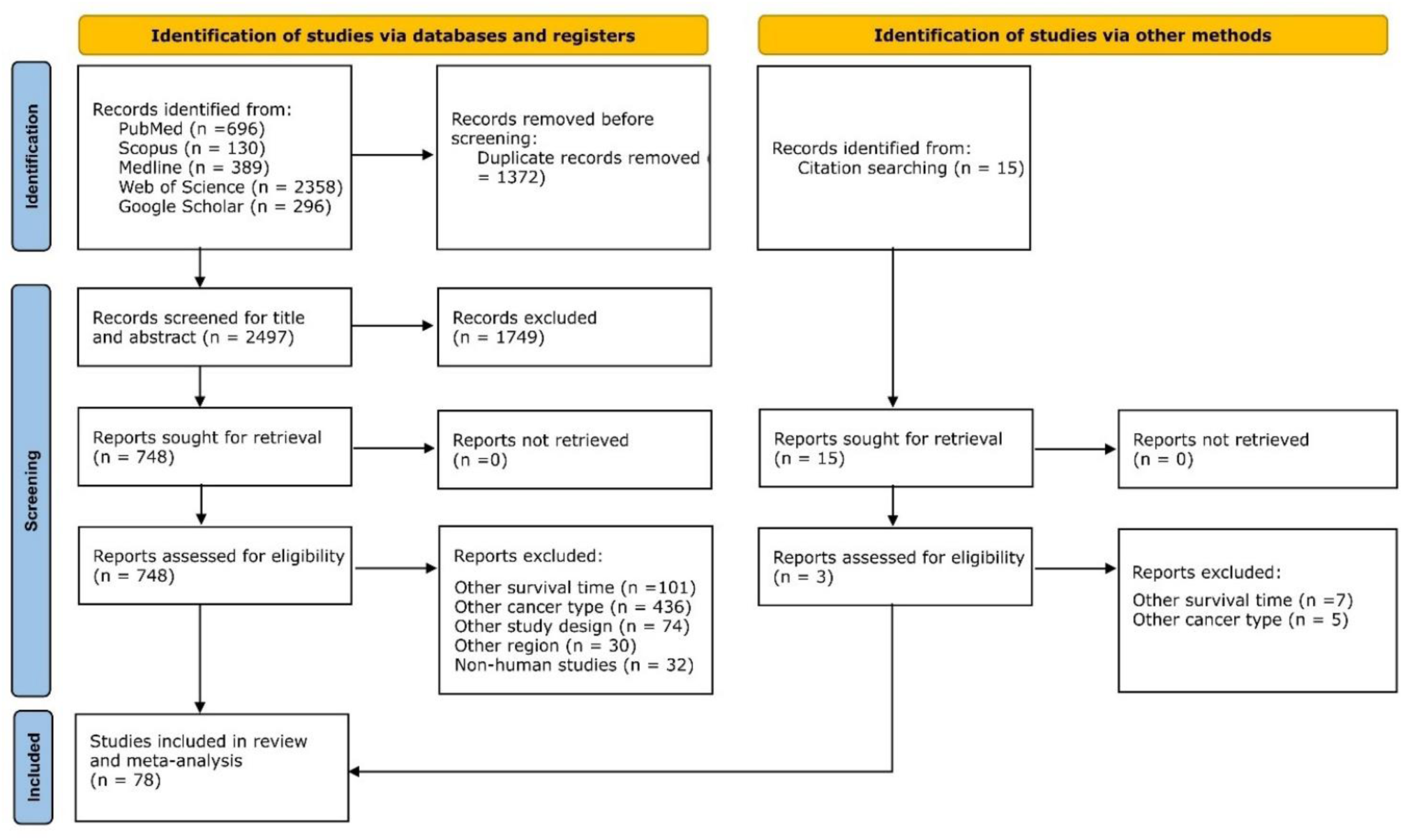
PRISMA flowchart. Flowchart showing the number of records identified, screened, and deemed eligible for pooled meta-analysis.

**Figure 2.**
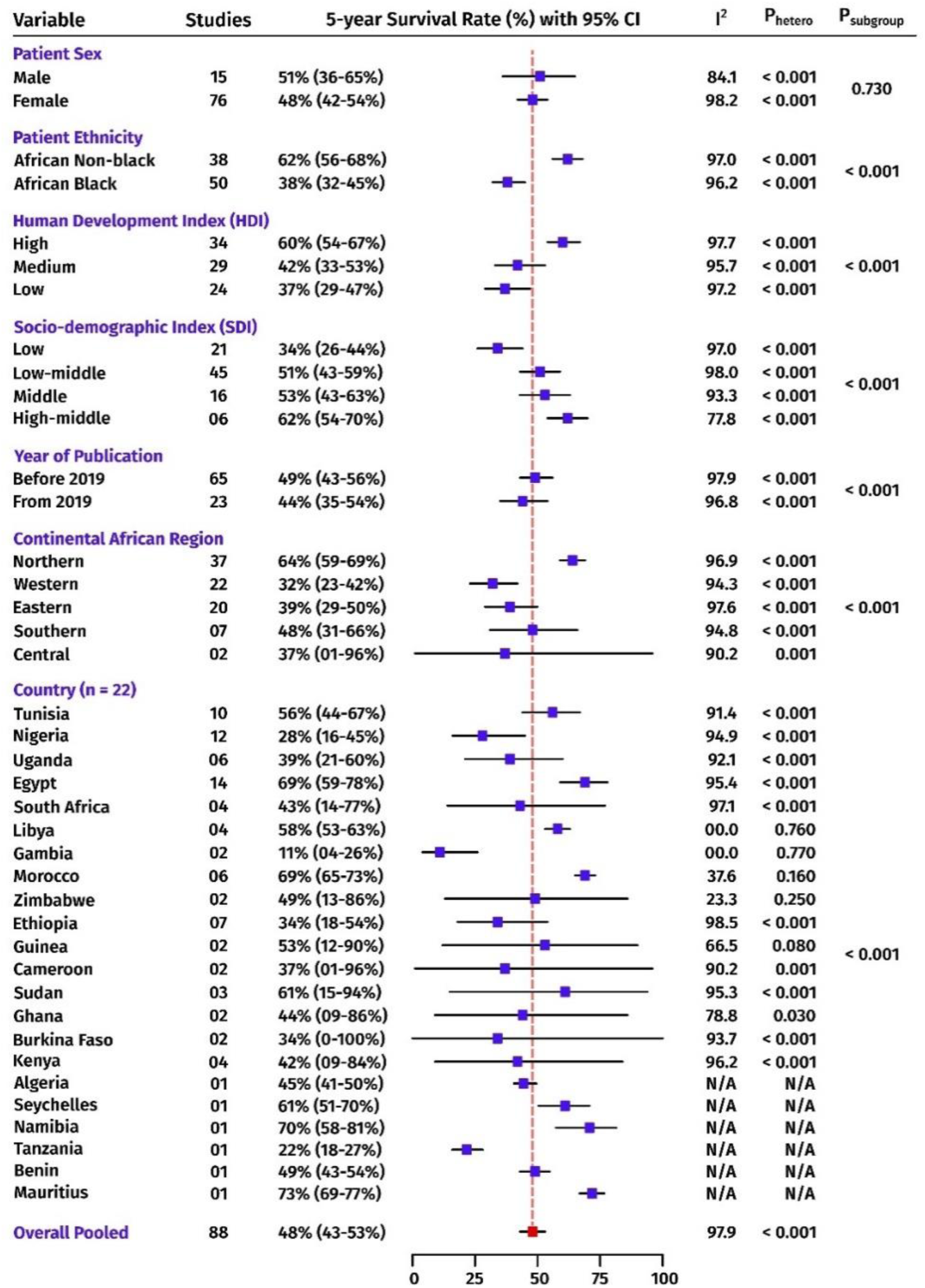
Sub-group meta-analysis for 5-year breast cancer survival rates across Africa. The blue box represents the overall survival rate of the respective sub-groups, while the red box indicates the pooled overall survival rate. The horizontal lines traversing each box depict the corresponding confidence intervals. P_hetero_ represents the P value for heterogeneity test while P_subgroup_ indicates the P value for the test for subgroup analysis. I^2^ represents the statistical heterogeneity (%) in the analysis. N/A – not applicable due to single study meta-analysis.

**Figure 3.**
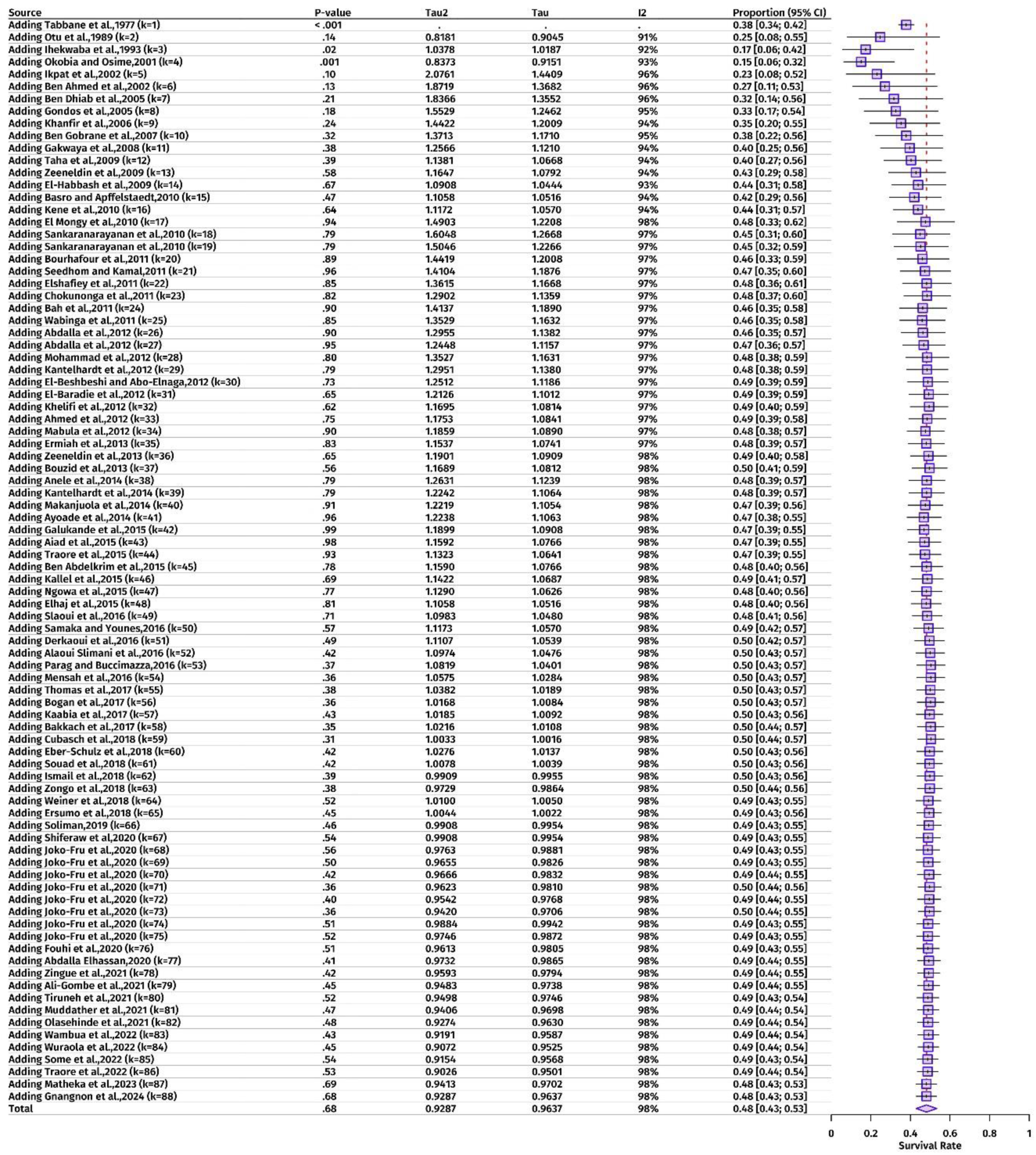
Cumulative meta-analysis for 5-year survival rates in Africa, adding studies in chronological order. The blue box represents the overall survival rate of the respective study, while the horizontal lines traversing each box depict the corresponding confidence intervals. Tau, Tau^2^, and I^2^ represent the different measures of statistical heterogeneity in the pooled estimate.

### 3.1. Meta-analysis

The overall pooled survival rate was 48% (95% CI: 43-53%), with high heterogeneity (I² = 97.9%), indicating substantial variability among the included studies. Sub-group-wise, a marginal discrepancy in survival rates was observed between the sexes, with females exhibiting a slightly lower survival rate (48%) compared to males (51%), though this difference was not statistically significant. However, a significant ethnic disparity was observed, with African non-black patients exhibiting a higher survival rate (62%) compared to African black patients (38%). Survival rates were also found to be significantly correlated with socioeconomic development. Regions exhibiting higher human development index (HDI) levels demonstrated superior outcomes, with high HDI regions exhibiting a 60% survival rate, medium HDI regions a 42% survival rate, and low HDI regions a 37% survival rate.

A similar pattern was observed with the Socio-demographic Index (SDI), where high-middle SDI regions exhibited the highest survival rate (62%) and low SDI regions exhibited the lowest (34%). Furthermore, it was observed that studies published prior to 2019 indicated a higher survival rate (49%) than those conducted from 2019 onwards (44%). Although the impact of the COVID-19 pandemic might be considered a contributing factor in this decline, we would urge caution in this interpretation. The reported studies were conducted retrospectively, and the year of publication may not be a reliable measure of the effect in question. This observed decline has been explored and addressed in greater detail later in the paper.

Furthermore, considerable variability was observed across African regions, with Northern Africa having the highest survival rate (64%) and Western Africa having the lowest (32%). Country-specific analyses indicate that Mauritius (73%) and Namibia (70%) recorded the highest survival rates, whereas the Gambia (11%) had the lowest rates.

We next conducted a cumulative chronological meta-analysis to examine the evolution and trajectory of 5-year survival rates. The analysis, which commenced with Tabbane et al., (1977) [68] and concluded with Gnangnon et al., (2024) [83], revealed interesting trends. The observed survival rates ranged from 15% to 50% between 1977 and 2024. The P values, which were calculated by comparing the observed effect with a previously reported null effect of 47%, exhibited considerable variation. Among the included studies, only three demonstrated statistically significant results (P < 0.05) from this null effect. As the number of studies increased, the overall survival rate rose and the confidence intervals narrowed, reflecting enhanced precision in the cumulative estimate. However, since the early 2010s, the overall survival rate has stabilized around 48-50%, with no discernible or predictable trend in the future.

### 3.2. Meta-regression

Meta-regression analyses were performed to evaluate the influence of different factors on 5-year survival rates. Among the continuous factors, mean years of schooling (MYS) and quality score (QS) of the included studies were both found to have significant influences (P = 0.001 and 0.023, respectively). MYS and QS accounted for 10.85% and 4.80% of the heterogeneity in survival, respectively. Each additional year of mean schooling was associated with a 2.41% (95% CI: 0.98-3.84%) increase in 5-year survival. Similarly, each 1-point increase in quality score was associated with a 1.39% (95% CI: 0.19-2.59%) increase in survival (**Figure 4**).

**Figure 4.**
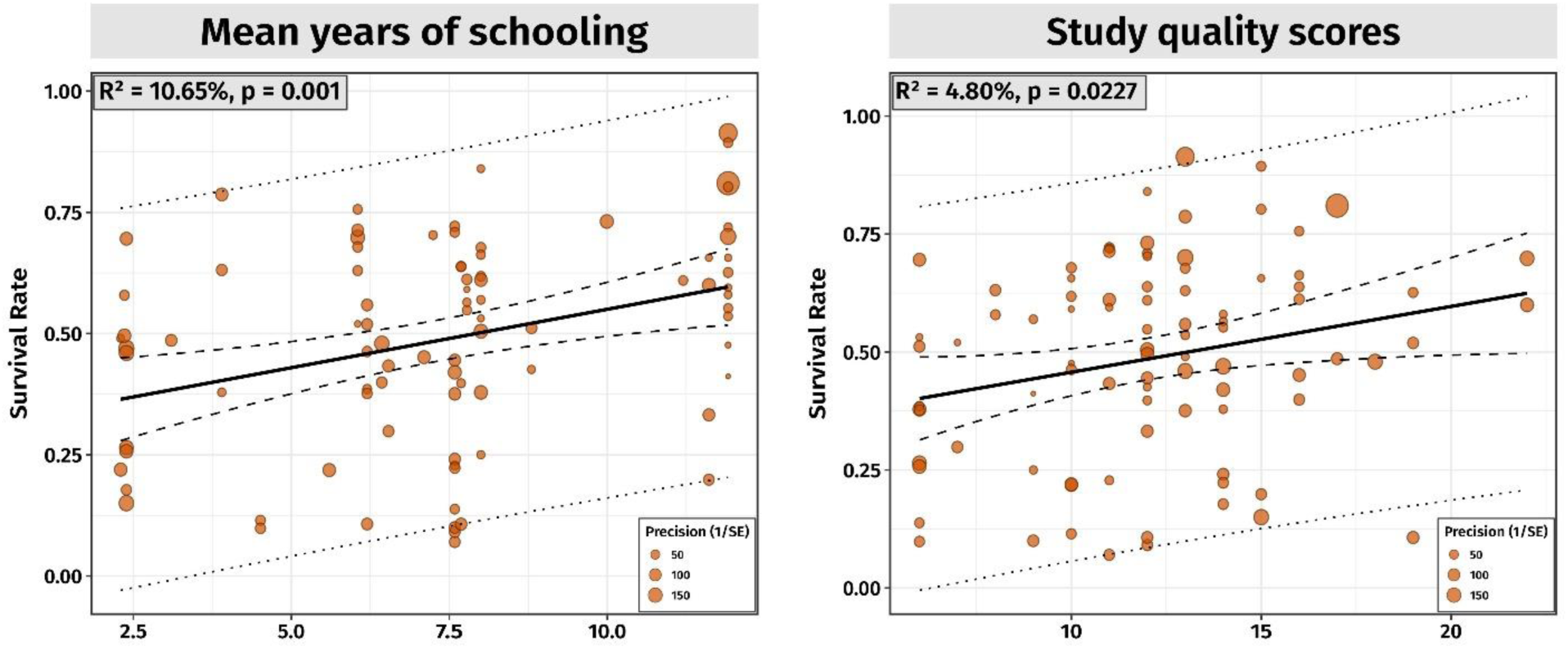
Bubble plots for the relationship between mean years of schooling (MYS) and quality score (QS) of the included studies with the 5-year survival rate of breast cancer patients. Each point represents a study, with the size of the points indicating the precision (1/standard error) of the data. The solid black line represents the regression line, the dashed lines indicate the 95% confidence intervals, and the dotted lines show the prediction intervals.

Additional analyses for categorical predictors revealed significant associations for survival rates with country, patient ethnicity, African continental regions, HDI quartiles, and SDI quintiles, indicating that survival rates are significantly influenced by these factors. However, no significant associations were found for patient sex and year of publication (**Figure 5**).

**Figure 5.**
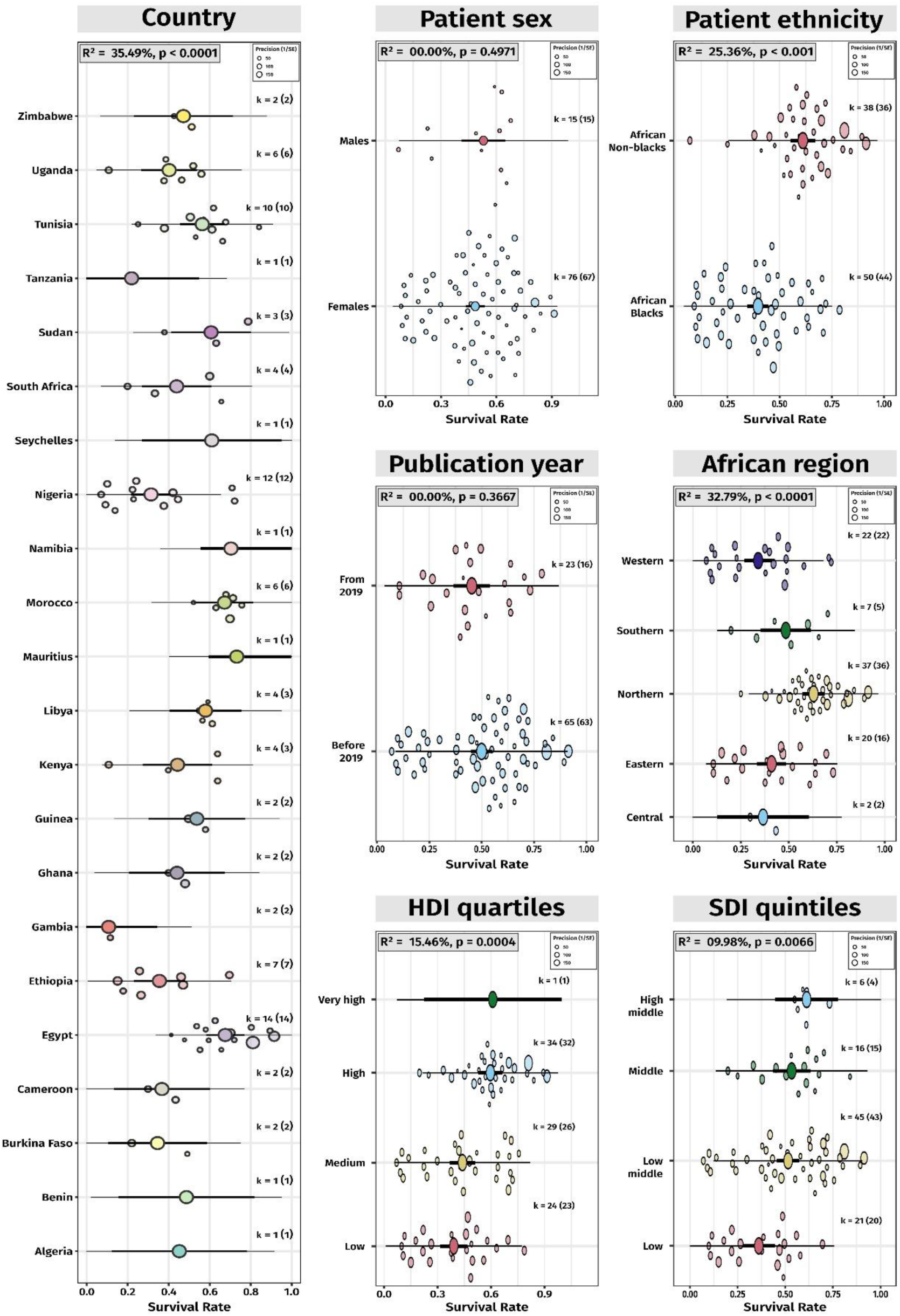
Orchard plots for the meta-regression analyses of the 5-year survival rate and categorical predictors. The plots show the pooled survival rate (trunk), 95% confidence intervals (branches), and prediction intervals (twigs). Bubble sizes indicate precision (1/standard error) of each study, with larger points representing higher precision.

### 3.3. Publication Bias and Small Study Effects

We used the Egger’s test to assess funnel plot asymmetry. The test result indicated significant asymmetry and was considered suggestive of potential publication bias (t =-3.15, P = 0.002. The bias estimate was-4.007 with a standard error (SE) of 1.271. Next, the trim-and-fill test was performed which included 114 studies, with 26 hypothetical studies added to adjust for bias (**Figure 6**). The adjusted 5-year survival rate using the random effects model was 62% (95% CI: 55-67%), with a prediction interval ranging from 10-96%. This adjustment indicates that the true effect size is likely to be within this broader range, accounting for potential missing studies.

**Figure 6.**
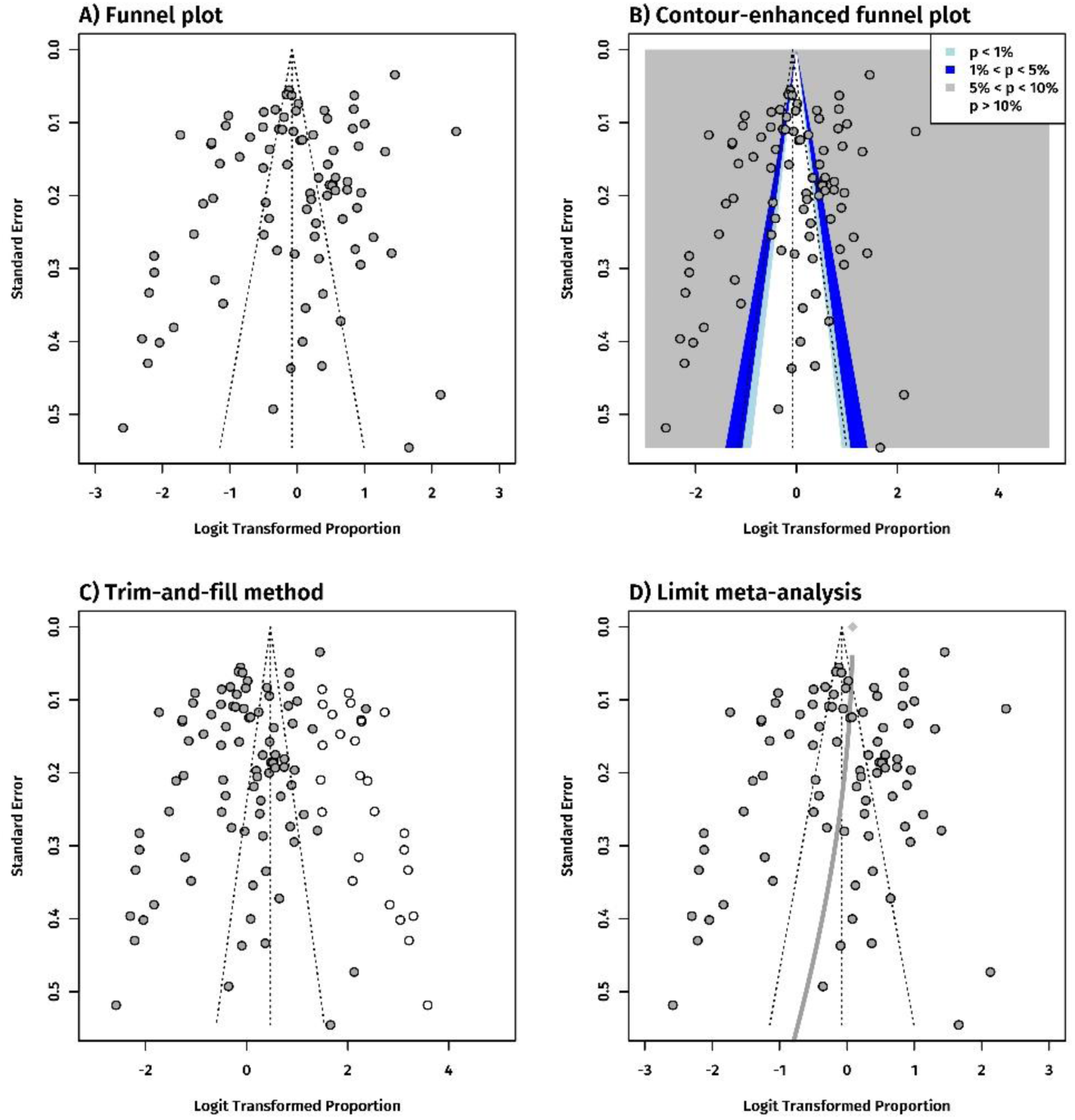
Funnel plots for assessing publication bias in studies reporting breast cancer survival rates. Standard funnel plot for 88 studies (panel A); contour-enhanced funnel plot for 88 studies (panel B); Trim-and-fill funnel plot with 26 hypothetical studies (white dots) added to 88 studies (gray dots; panel C); and Limit meta-analysis funnel plot showing increasing bias with standard error. Gray diamond shows the adjusted average effect when standard error is zero.

The analysis still quantified substantial heterogeneity among the included studies, with a tau-square (τ^2^) value of 1.837 (95% CI: 1.446-2.509). The I^2^ statistic was observed to be extremely high at 98.4% (95% CI: 98.3-98.5%), indicating that nearly all the variability in effect estimates is due to heterogeneity rather than random chance. The limit meta-analysis, on the other hand, provided a 5-year survival estimate of 52% (95% CI: 45-59%).

Heterogeneity statistics indicated a τ^2^ value of 0.9287, with an I² of 97.9% (95% CI: 97.7-98.1%), reflecting substantial heterogeneity among the studies. Additionally, the test for small-study effects demonstrated Q-Q’ of 430.25 (P < 0.0001), suggesting the presence of small-study effects. The residual heterogeneity test produced Q’ = 3719.79 (P < 0.0001), indicating significant residual heterogeneity beyond small-study effects.

### 3.4. Comparison with 2018 African Estimates

A previous meta-analysis, which included literature up to October 2018, identified 54 eligible records from 14 countries [10]. The authors reported a pooled overall 5-year survival rate of 53% (95% CI: 46-60%), while our own findings indicated an estimate of 48% (95% CI: 43-53%). We conducted a one-sample Z-test for proportions to compare the two estimates and found no statistical significance (P = 0.071). Nonetheless, given that our estimates indicated a lower survival rate than previously reported, we explored the cause for this observation.

At first, we postulated that our larger and more diverse data set, comprising 22 countries, could have yielded a more precise estimate. To explore this, we subset our data into two groups, one part with data from same 14 countries as reported from 2018 meta-analysis and the other part with data from 8 new countries (**Figure 7**). The forest plot demonstrated that the estimate derived from the former group remained inferior to the estimates reported by the 2018 study (47% vs 53%).

**Figure 7.**
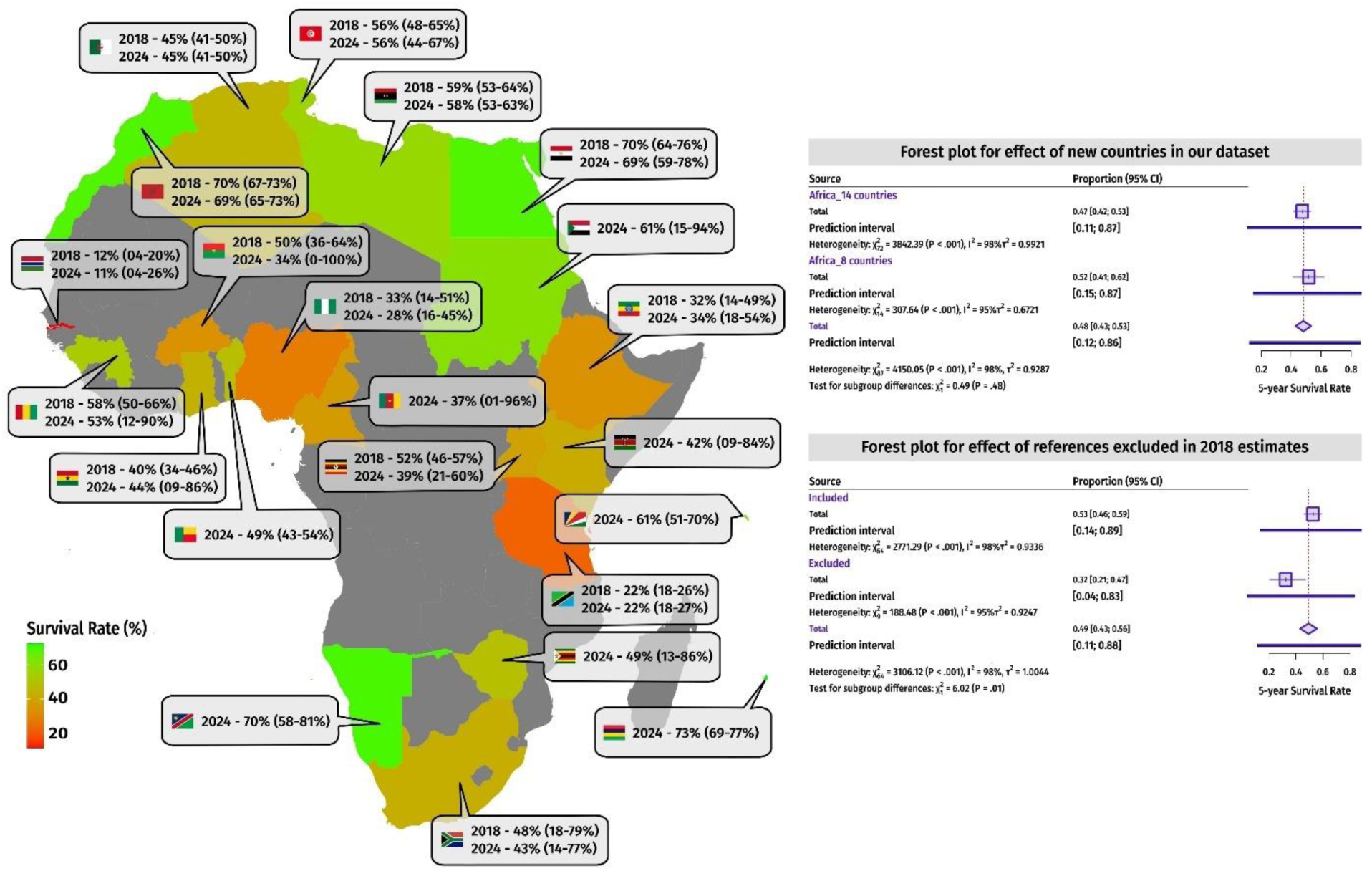
Comparison of our current 5-year survival estimates with the previous 2018 estimates. A territorial map of continental Africa is shown (on the left) with 5-year survival estimates and 95% confidence interval from 2018 and 2024. The survival estimates have been rounded off to the nearest whole number. Two forest plots are shown (on the right) that were used to estimate the effect of addition of eight new countries in our 2024 estimates and the effect of missed/excluded references in the 2018 estimates.

Subsequently, we cross-referenced the meta-analyzed studies from the previous work and those from the present study. We observed that the authors had not included 10 studies that were published before 2018 and met their inclusion criteria, potentially attributable to the limited number of databases they searched (Medline, Embase, and Cochrane Library). These studies were from Uganda, Gambia, Ghana, Ethiopia, Nigeria, Cameroon, Sudan, and Zimbabwe. Inclusion of these 10 studies yielded an estimated 5-year survival rate of 49% (95% CI: 43-56%; **Figure 7**), which is in close alignment with our pooled overall survival rate of 48%.

This decrease observed in 2018 estimates was expected since the studies not included were primarily from low and low-middle SDI countries. In our view, these findings have two broader public health ramifications. First, although not significantly different from our estimates, the previously reported survival estimates appear to be inflated due to incomplete catch from the investigated biomedical databases, highlighting the need for a comprehensive search strategy. Second, as observed with our cumulative meta-analysis, the 5-year survival rate has somewhat stagnated in Africa. Apart from Ghana, all other countries either observed a modest decrease or no change in 5-year survival estimates (**Figure 7**). Burkina Faso and Uganda experienced the largest declines, although the one-sample z-test for proportions was not significant (P = 0.268 and 0.195, respectively).

### 3.5. Comparison with Global Estimates among Female Patients

We also conducted a generic Google Scholar and PubMed search to look for pooled 5- year survival rates for breast cancer patients. The search was specifically designed to retrieve systematic reviews and meta-analyses published in the last decade that included data from multiple countries and had pooled the results based on either regional grouping, WHO regions, or at the continent level. Consequently, three such studies were identified for comparison [97–99]. Given that these studies reported survival rates exclusively for female patients, they were compared with our estimates derived from the female subset only (**Figure 8**).

**Figure 8.**
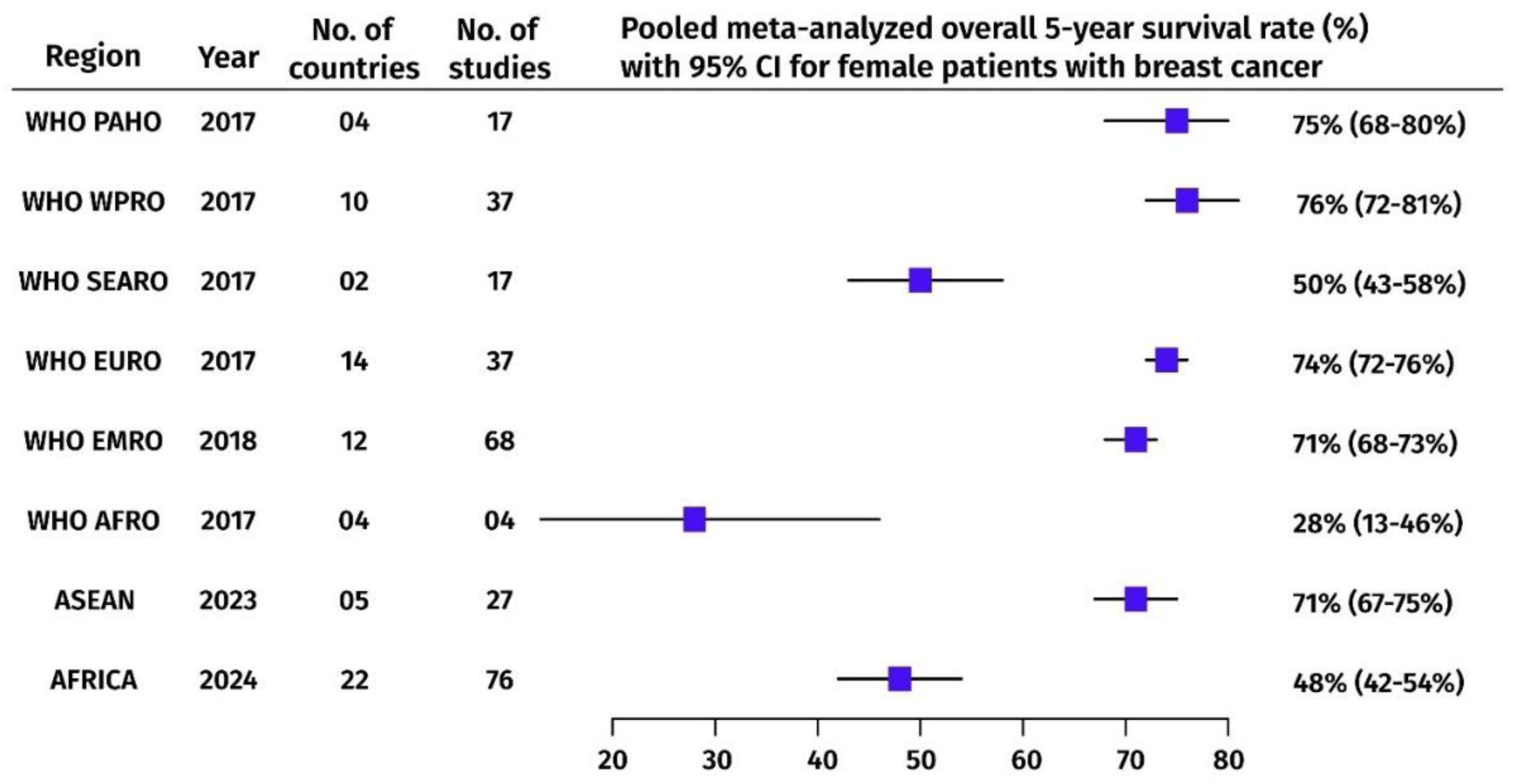
Comparison of our current 5-year survival estimates with the previously reported global estimates. Forest plot showing the pooled meta-analyzed overall 5-year survival rate with 95% confidence interval for female patients with breast cancer across different regions. WHO – World Health Organization; PAHO – Pan American Health Organization; WPRO – Western Pacific Region; SEARO – South-East Asia Region; EURO – European Region; EMRO – Eastern Mediterranean Region; AFRO – African Region; ASEAN – Association of Southeast Asian Nations.

A comparative analysis of five-year survival rates for female patients across different regions revealed significant differences between our 2024 estimates and previously reported estimates from all regions (one-sample Z test P < 0.001), except for the WHO SEARO region (one-sample Z test P = 0.469). In 2017, the WHO WPRO and PAHO regions recorded the highest global survival rates of 76% and 75%, respectively. Similarly, the WHO EURO and WHO EMRO regions reported high survival rates of 74% and 71%, respectively.

Although the survival rate in the WHO SEARO region was slightly higher than our 2024 estimates, there is clearly room for improvement in both regions. Notably, the estimates from the WHO AFRO region, based on just four studies, reported a much lower survival rate of 28% in 2017. Upon subsetting and analyzing the results for female patients from WHO AFRO countries (48 studies; 17 countries), we observed a 5-year survival rate of 38% (95% CI: 32-44%). This indicates a considerable improvement in the WHO AFRO region’s survival rate by 2024. Despite this progress, the survival rate remains substantially lower than in other WHO regions.

## 4. Discussion

Our meta-analysis of 5-year survival rates for breast cancer patients in Africa offers comprehensive insights into the current state of clinical outcomes across the continent. We underscore the considerable disparities in survival rates across different regions and socio-economic contexts. The overall survival rate of 48% in Africa is markedly lower than those reported in high-income countries, where survival rates frequently exceed 70-80% [97–99].

The sex-stratified analysis indicated no differences in survival rates between male and female patients, suggesting that both genders encounter considerable challenges in the context of early diagnostics and timely and effective treatment. In their study, Gómez-Raposo et al., found that when age is accounted, breast cancer survival rates are similar for both males and females [100]. Others have also reported similar findings when the cohorts were age-, region-, stage-, and/or propensity-matched [101–104]. Nonetheless, the marginally higher survival rate observed in males is interesting, as literature seems to suggest a worse prognosis for male breast cancer patients. The lower survival rate observed in males has been suggested due to males having a greater likelihood of being diagnosed at an older age, a higher prevalence of advanced disease stages, and a tendency to adhere less strictly to treatment regimens [105]. Furthermore, the issue is compounded by a lack of awareness and the absence of targeted screening programs for male patients.

In the context of our study, data were available for male patients only from nine countries in Africa. The rarity of male breast cancer surveillance and reporting may have resulted in the inclusion of smaller, non-representative cohorts, thereby influencing the observed outcomes. Furthermore, our methodological approach of categorizing studies based on a threshold criterion (≥80% of one sex) may also contribute to this observation. Finally, it may also be due to the absence of triple-negative cases in the male cohort, suggesting that certain aggressive subtypes are less common among African men [106]. This absence of aggressive difficult-to-manage phenotypes could influence and contribute to the observed higher survival rates.

Furthermore, African black women are more likely to be diagnosed with aggressive subtypes of breast cancer, such as triple-negative breast cancer (TNBC) [105,107]. TNBC is characterized by the absence of estrogen, progesterone, and HER2 receptors, which renders it more challenging to treat and is associated with a poorer prognosis [108–109]. The *ANKLE1* gene, which confers protection against TNBC, was found to be less prevalent in African American women compared to white women of European descent [110]. Moreover, African American women with a mutation in the Duffy (*DARC*) gene, which is involved in the inflammatory response, have an elevated risk of developing TNBC [110].

Northern Africa, with its relatively high survival rates, benefits from a more robust healthcare infrastructure, more efficacious screening programs, and greater public awareness about breast cancer. It is frequently the case that countries in this region are ranked high on HDI and SDI evaluations [12–13]. Conversely, the lowest survival rates are observed in Western Africa, which highlights the necessity for significant investments in healthcare services and resource allocation. A country-specific analysis revealed significant contrasts, with Mauritius recording the highest survival rate and Gambia having the lowest. These discrepancies may be attributable to discrepancies in healthcare systems, the availability of cancer treatment facilities, and national and international health partnerships and policies.

For example, in Mauritius, healthcare is regarded as a fundamental human right, with free public health services for primary care being made available. Every household is situated within three miles of a primary care provider, thereby ensuring accessible medical care for all. Active investments are being made by the Ministry of Health and Wellness to construct a national cancer hospital, while upgrading facilities at regional hospitals to ensure that cancer treatment is provided free of charge to all citizens [111]. In contrast, the availability of facilities for breast cancer screening, diagnosis, and treatment in Gambia is limited. Many of the diagnostic and care centers are located at distances ranging from 10 to 45 kilometers from patients’ residences. Consequently, nearly half of the Gambian population lacks access to pathologic diagnosis and surgical management of breast cancer within these distance thresholds [112]. In other countries with high overall breast cancer survival rates, such as Seychelles and Namibia, which have higher HDI values, the density of healthcare professionals (HCPs) per 1,000 population exceeds the United Nations Sustainable Development Goals (SDG) density target of 4.45 HCPs per 1,000 population. Furthermore, these countries maintain healthcare quality ratings above 80% [113].

A comparison of the 2024 estimates with the adjusted 2018 estimate and the findings from our cumulative meta-analysis indicates a stagnation of survival rates around 48-49% over time. This may be attributed to several factors. The healthcare infrastructure across many African countries remains underdeveloped, with the availability of essential diagnostic tools and advanced treatment options remaining constrained, particularly in rural and low-resource settings [114]. This inadequacy results in delayed diagnoses and suboptimal treatment outcomes. Socioeconomic and cultural mistrust in Western medicine, in conjunction with pervasive poverty and economic disparities, impede the capacity of patients to access essential healthcare services at a reasonable cost [115–116]. Traditional beliefs and stigma associated with cancer can act as a deterrent for individuals seeking medical care [117–118]. Despite the importance of efforts to raise awareness and educate the population about the significance of early detection and treatment, initiatives have thus far proven inadequate in terms of both scope and impact.

Systemic issues, including political instability and corruption, serve to further exacerbate these challenges, impeding the implementation of cancer care programs [119]. Ultimately, inadequate financial resources have an adverse ripple effect throughout the healthcare system, from the unavailability of essential medications to the lack of sufficient training for HCPs, perpetuating a vicious cycle of inadequate care and poor patient outcomes. Our observations also highlight that sustained improvements in particular regions or countries are insufficient to significantly alter the overall continental survival rate. The considerable heterogeneity observed in the studies highlights the uneven distribution of healthcare resources and the variable quality of care across different countries and regions.

Another notable finding was the positive correlation between mean years of schooling (MYS) and breast cancer survival rates. Education plays a crucial role in enhancing health outcomes by fostering health literacy, which provides for greater awareness of breast cancer symptoms, the significance of early detection, and adherence to treatment regimens. Higher levels of educational attainment afford patients the ability to navigate the healthcare system with greater confidence and to make more informed decisions regarding their care. Furthermore, the analysis underscores the influence of study quality on the reported survival rates, indicating that higher-quality studies are associated with more favorable survival outcomes. This highlights the necessity of rigorous study design and adherence to methodological standards to produce reliable and accurate data. The quality of the studies included in the meta-analysis is reflective of the variability in healthcare infrastructure, research funding, and expertise across different regions.

It is hence necessary to improve the caliber of research conducted in resource-limited settings to generate evidence-based unbiased data and subsequently inform policy and practice. The observed heterogeneity in survival rates is significantly influenced by region, country, race, SDI, and HDI. This indicates that geographical and socio-economic factors exert a more pronounced influence on breast cancer outcomes in Africa than temporal trends or gender or ethnicity. The evidence of significant publication bias, as indicated by Egger’s test, suggests that the reported survival rates may be overestimated due to the under-reporting of smaller studies with less favorable outcomes.

### 4.1. Limitations

While we followed a comprehensive protocol, there are several methodological limitations that warrant consideration. The substantial heterogeneity among the included studies indicates significant variability in survival rates, likely due to differences in study design, population characteristics, and healthcare systems, complicating the generalization of results across the continent. The presence of publication bias suggests that smaller studies might be under-represented, potentially skewing the overall survival rate estimate despite adjustments using the trim and fill method. Additionally, the analysis included studies from only 22 out of 54 African countries, limiting the representativeness of the findings and potentially overlooking variations in survival rates in countries with no available data.

The reliance on published studies means that the quality and accuracy of the data are contingent on the methodologies and reporting standards of the original research, introducing inconsistencies. For instance, our approach for classification of studies based on patient sex does not fully account for the nuanced effects of sex on breast cancer outcomes and could lead to an over-estimation or under-estimation of survival rates for each sex. Additionally, while this threshold ensures the majority sex group’s characteristics predominantly drive the outcomes, it highlights the necessity for further research to comprehensively disaggregate, record, and report data by sex. Another crucial disadvantage of reporting inconsistencies is that tumor characteristics (staging, TNM classification, histological classification, localization, primary tumor/metastasis) and clinically relevant parameters (patient age, treatment modality, follow-up duration, quality of life outcomes) cannot be compared and pooled together.

Such limitations have been reported by previous global meta-analytical studies as well where authors reported that data from low-and middle-income countries such as those in Africa and Asia had >50% of patients with unknown breast cancer stages to report [6]. Furthermore, assessment of tumor staging should be done using both clinical and radiological methods. However, a meta-analysis for Sub-Saharan African countries noted that < 25% of the studies reported the staging methods used with a few studies relying only on clinical assessment for staging [120]. Finally, socio-economic factors such as income and access to healthcare, which significantly impact patient outcomes, were not fully accounted for in the analysis. Addressing these limitations in future studies will enhance the accuracy and reliability of the findings, contributing to better-informed healthcare policies and interventions.

### 4.2. Recommendations based on current estimates

We reiterate the longstanding and urgent necessity of implementing targeted healthcare policies to address the considerable regional disparities that are prevalent among African countries. The observed correlation between higher survival rates and countries with a high HDI, MYS, and SDI underscores the pivotal role of robust healthcare infrastructure, education, and socio-economic development in enhancing cancer care. It is incumbent upon policymakers to prioritize the allocation of resources to regions with the lowest survival rates, such as Western Africa, and to invest in the development of healthcare facilities, diagnostic tools, and treatment centers. Furthermore, it is crucial to enhance the training of HCPs in oncology, molecular diagnostics, and palliative care. Implementing culturally sensitive public health campaigns, adopting universal healthcare, and expanding insurance coverage to include cancer treatment are essential steps.

The considerable heterogeneity and publication bias identified in our meta-analysis underscores the need for more comprehensive and high-quality record keeping and research, including the establishment of national cancer registries. International collaboration and partnerships with global health organizations can facilitate the transfer of knowledge and the allocation of funding for improvements in cancer care. There is a need to develop and implement an Africa-wide public health framework for the development of targeted strategies that address the specific needs and challenges of different regions and populations in Africa.

## 5. Conclusions

Our findings highlight the significant regional disparities in breast cancer survival rates across the African continent, reinforcing the urgent and sustained need for targeted health interventions. The substantial heterogeneity among studies reflects the variability in the quality of and access to health care across countries. The higher survival rates in countries with high HDI and SDI values underscore the importance of robust health systems and socioeconomic development in improving survival. Gender differences in survival were found not to be a covariate, highlighting the importance of controlling for other clinical and socio-demographic factors. These findings provide a roadmap for developing effective cancer screening programs and health policies, including implementation of uniform reporting standards and establishment of national cancer registries, to improve breast cancer surveillance, monitoring, and survival rates across the continent.

## 6. Declarations

a. **Conflicts of interest** – None to declare.
b. **Funding** – None to declare.
c. **Ethical approval** – Not applicable.
d. **Data Availability** – All data generated is presented in the manuscript and appendices. Clarifications on methodology and requests for the R code can be directed to the corresponding authors.
e. **Informed Consent** – Not applicable.
f. **Acknowledgements** – None to declare.
g. **Author contributions** – ABP and NJ conceptualized the present study. Data curation, collection, and screening was done by ABP, EKK, AGB, JK, AAE, BS, IN, and BOO with inputs and validation from NJ. Methodology, investigations, visualizations, and formal analysis were done by EKK and NJ. Software and code were maintained by EKK and NJ. Project administration and supervision was done by ABP and NJ. Data validation was done by EKK and NJ. The original draft was prepared by EKK, JK, ABP, and NJ while all authors were involved in revising the manuscript. All authors have read the manuscript and agreed on its contents for publication.

## Data Availability

All data generated is presented in the manuscript and appendices. Clarifications on methodology and requests for the R code can be directed to the corresponding authors.

## Notes

### Competing Interest Statement

The authors have declared no competing interest.

### Clinical Trial

Not applicable.

